# New reproduction numbers for the visible and real epidemic dynamics

**DOI:** 10.1101/2025.01.10.25320319

**Authors:** Igor Nesteruk

## Abstract

Known basic and effective reproduction numbers are based on registered (visible) cases despite that asymptomatic and unregistered patients are characteristic of almost all infection diseases. New reproduction numbers - the ratios of the real numbers of infectious persons (symptomatic and asymptomatic) at different moments of time and a simple method of their estimation with the use of visible (registered) cases only were proposed. The novel general SIR (susceptible-infectious-removed) model was used and the results of calculations for the pertussis epidemic in England are presented. New approach could help to control different epidemics, in particular new waves of COVID-19.

---

Known basic *R*_*0*_ and effective *R*_*t*_ reproduction numbers [1, 2] and methods of their estimation [2-9] use the registered (visible) cases *V*^*(v)*^. Nevertheless, asymptomatic and unregistered cases are characteristic of almost all infections including SARS‐CoV‐2 [10-15] and pertussis [16]. Moreover, it is difficult to estimate the numbers of infectious persons *I(t)* (depending on time *t*) with the use of *V*^*(v)*^*(t)* even for the totally visible epidemics [2-9] (when the ratio of real infections to the registered ones *β* =1). In this study we propose new reproduction numbers - the ratios of the numbers of infectious persons at different moments of time – as important characteristics of epidemic dynamics and a simple method of their estimation with the use of visible (registered) cases *V*^*(v)*^*(t)* only and the novel general SIR (susceptible-infectious-removed) model [17].

For simplicity let us use the new reproduction number in the following form:

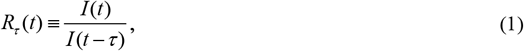

which shows the rate of increasing (decreasing when *R*_*τ*_ (*t*) <1) the real numbers of infectious persons during time *τ*. *R*_*τ*_ (*t*) *=*1 corresponds to the equilibrium (endemic) state. The novel general SIR model relates the numbers of susceptible persons *S(t)*, visible (symptomatic) *I*^*(v)*^*(t)* and hidden *I*^*(h)*^*(t)* (asymptomatic) patients (*I(t)*=*I*^*(v)*^*(t)+I*^*(h)*^*(t))*, and corresponding removed persons *R*^*(v)*^*(t)* and *R*^*(h)*^*(t)* (see[17] or Supplementary **S1**). Supposing all parameters of the model to be constant during the *i-th* epidemic wave, a simple relationship

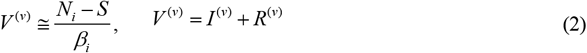

can be obtained (*V*^*(v)*^*(t)* is accumulated number of visible cases; *β*_*i*_ is the visibility coefficient; *N*_*i*_ is the number of susceptible persons before the outbreak of the *i-th* epidemic wave, the first eq. (2) holds exactly for cases, when all patients are removed before the outbreak of the the *i-th* epidemic wave, [17], **S1**). Taking into account (2) and the differential equation:

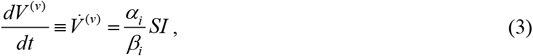

(*α*_*i*_ is infection rate, [17], (S9)), the following relationship can be obtained:

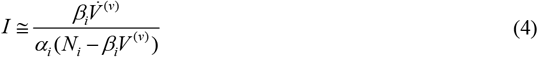

Substitution of (4) into (1) yields

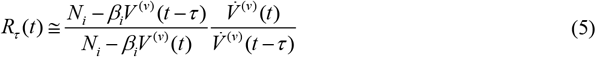

If *N*_*i*_ is much lager than the real number of cases (approximately equal to *β*_*i*_*V*^*(v)*^, [17]), eq. (5) yields :

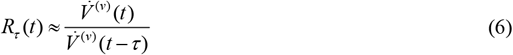

At discrete moments of time *t*_*j*_ and for *τ =t*_*j*_ *–t*_*j-1*_

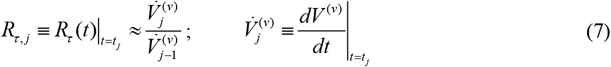

Since the derivatives in (6) and (7) are theoretical estimations of the daily, monthly or weekly (depending on the unit of time) numbers of new cases, they can be estimated numerically as follows, [18]:

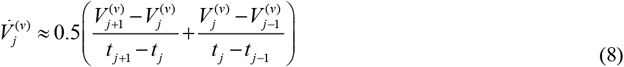

where 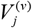 are the accumulated numbers of visible cases registered at moments *t*_*j*_. When the daily numbers of new cases are very random and show some weekly periodicity (e.g., during the COVID-19 pandemic), the smoothed characteristics [8, 19]:

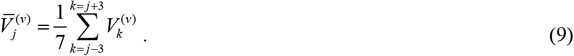

must be used in (8).

Thus, equations (6)-(9) allow estimating the rate of increasing the real numbers of infectious persons *R*_*τ*_ (*t*) with the use of observations of visible (symptomatic) cases only. An example of calculations for the pertussis epidemic in England in 2023 and 2024 [20] is shown in Table 1 (the last column) and in Fig. 1 (“crosses”). For example, the first value in the last column means that during the March 2023, the number of infectious (both symptomatic and asymptomatic) has increased by 48.7%. The accuracy of this and two next values *R*_*τ*,4_ and *R*_*τ*,5_ is limited, since the figures corresponding the transition from the endemic state in January and February 2023 (with constant monthly numbers of new cases, see the third column of Table 1) were used for their calculations (estimations (2)-(7) are valid for the constant values of the model parameters only). The accuracy of *R*_*τ, j*_; *j* = 9,10,11,12,13 is also limited due to the transition from the first to the second epidemic wave, **S2**). Other *R*^*τ, j*^ values agree quite well with the results of SIR simulations for *τ* =30.42 days (average duration of a month) both for the first and second epidemic waves (blue and red solid curves, respectively) despite the large differences between *t*_*j*_ and *t*_*j-1*_ values, which decrease the accuracy of numerical differentiation (8). It would be useful to compare weekly estimations, which could ensure better accuracy of (8) and shorter transition periods. Corresponding theoretical dashed curves are shown in Fig. 1. Dotted curves represent the SIR estimations of the effective reproduction numbers *R*_*t*_*(t)* (eq. (S11)). Almost critical values of the effective reproduction numbers (*R*_*t*1_ (*t*) ≈ *R*_*t*2_ (*t*) ≈ 1) and growing *R*_*τ*_ (*R*_*τ*, 21_ > *R*_*τ*, 20_) increase the probability of a new pertussis wave.

**Table 1.**
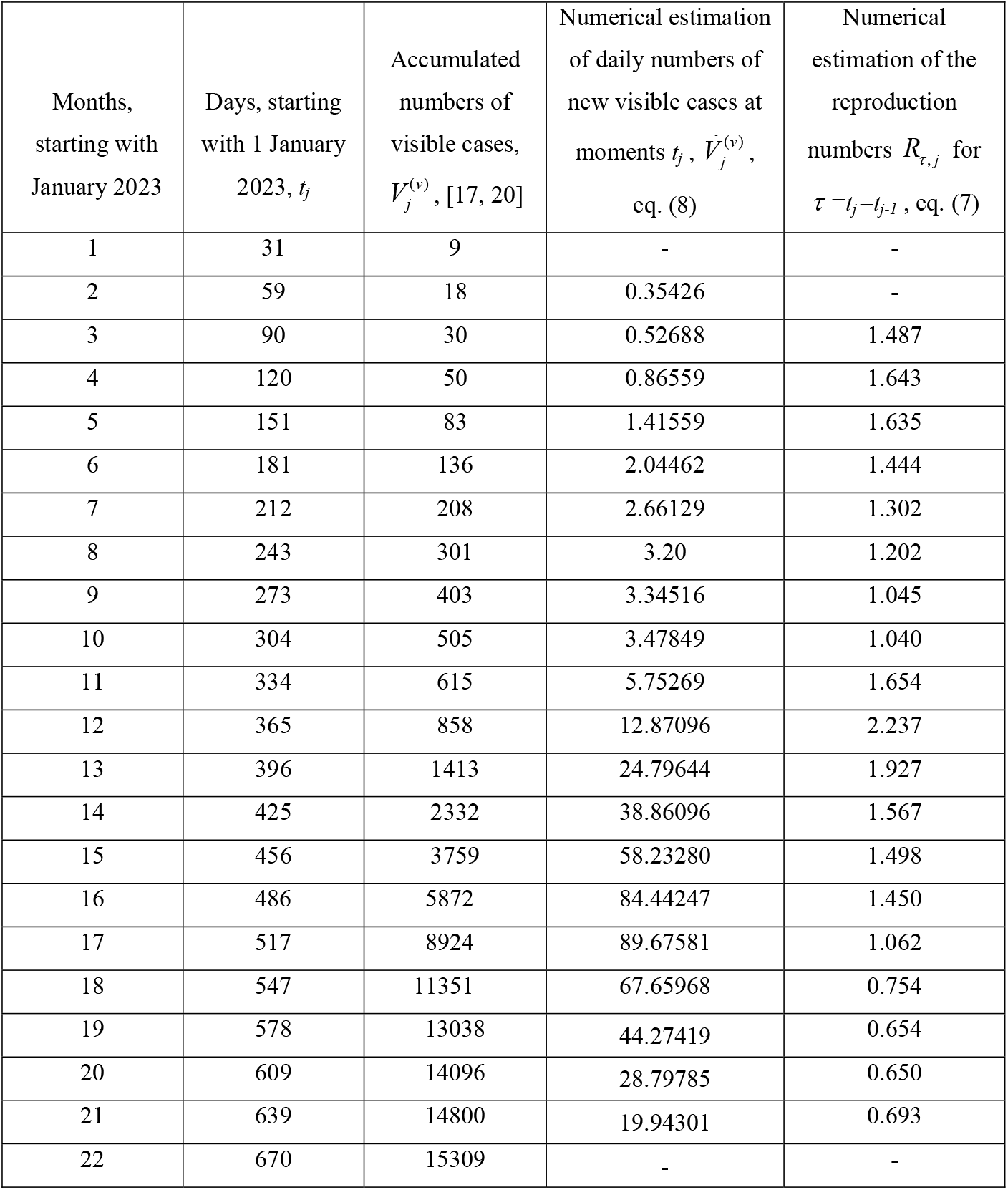
Accumulated numbers of confirmed pertussis cases in England in 2023 and 2024, estimations of the average daily numbers of visible cases and the reproduction numbers *R*_*τ*_ (*t*)

**Fig 1.**
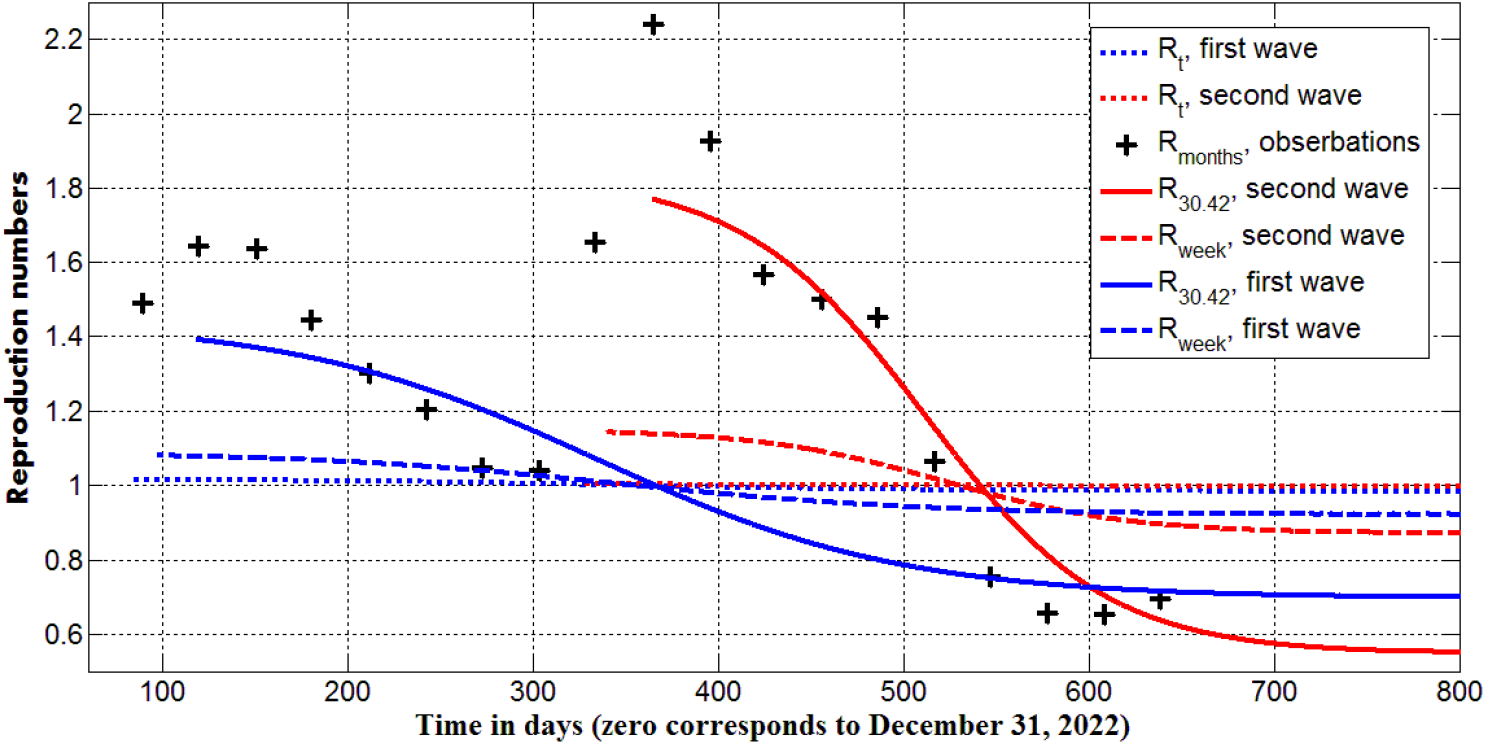
New reproduction numbers for the pertussis epidemic in England (“crosses”, Table1) and theoretical estimations for the fist (blue curves) and second (red curves) waves (S2). SIR simulations of *R*_*τ*_ (*t*) for two values of *τ* (*τ* =30.42 days, average duration of a month, solid curves; *τ* =7 days, dashed curves) and the effective reproduction numbers *R*_*t*_*(t)* (eq. (S11), dotted curves).

It follows from (1) and (7) that maxima of the numbers infectious persons and averaged daily numbers of new cases are reached at close moments of time. Since the presented estimations are valid at arbitrary values of the visibility coefficient *β* we can check this fact for numerous SIR simulations of COVID-19 dynamics at *β* =1, [8, 19, 21-23] and attempts to calculate the real numbers of cases and infectious [14, 15, 24]. At *β* =1, the analytical solution of SIR equations shows that the daily number of new cases start to decline before the maximum number of the infectious persons is achieved. This time difference can be calculated after identifications of the model parameters, but it is small according to the calculations and comparisons with the observations [8, 14, 15, 19, 21-24].

The new reproduction numbers allow estimating the changes in the concentration of infectious persons in the population of the volume *N*_*pop*_ (or probability to face a spreading patient *p*(*t*) ≡ *I* (*t*) / *N*_*pop*_, [19]). In particular, if a significant number of people move from a more infected region *A* to a less infected one *B* (*p*_*A*_ > *p*_*B*_), the increase of *R*_*τ*_ (*t*) (or average daily numbers of new cases, see (6)) occurs in region *B*. Such situation happened after the Russian full-scale invasion in Ukraine in February 2022. Millions of Ukrainian refuges [9, 25, 26] moved from the country with approximately 4 times higher values of *p*_*A*_ than in the whole world [9, 23] and caused the increase in the daily numbers of new COVID-19 cases in Germany, the UK and the whole world after 24 February 2022, [9]. These consequences of the aggression and the amount of compensation from Russia should be assessed.

The use of relationships (1), (7)-(9) can help to control the recent COVID-19 outbreaks, which are still dangerous [27-29]. Monitoring the average daily figures of new cases (with the use of (8),(9)) allows detecting the moments when the real numbers of infectious (visible and hidden) start to increase (according to (1) and (7)). Simultaneous increase in the number of tests (maintaining the high values of the tests per case ratio) could help to control the new epidemic waves completely (such examples demonstrated Australia, New Zealand, South Korea, Hong Kong, and Japan, [30])

## Clarification point

No humans or human data was used during this study

## Data Availability

All data produced in the present work are contained in the manuscript

## Acknowledgements

The author is grateful to Ulrike Tillmann, James Robinson, Robin Thompson, Matt Keeling, Paul Brown, and Oleksii Rodionov for their support and providing very useful information. This paper was written with the support of the INI-LMS Solidarity Programme at the University of Warwick, UK.

## Supplementary

### S1. The general model of visible and hidden epidemic dynamics and examples of analytical solutions

For every epidemic wave *i*, the compartment of infectious persons *I(t)* can be divided into visible (registered) and hidden (invisible/asymptomatic and unregistered) parts *I* = *I*^(*v*)^ + *I*^(*h*)^ appearing according to the visibility coefficient *βi* ≥ 1 and removing with rates 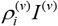 and 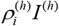. Then the general SIR model [8, 19] takes the following form [17]:

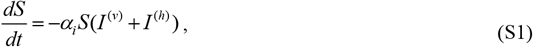

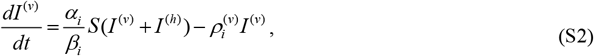

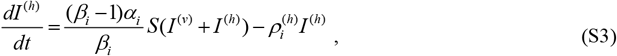

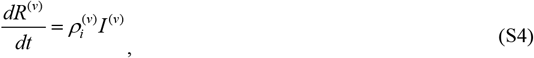

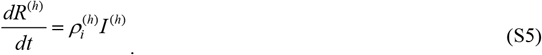

The compartment of removed persons *R(t)* is also divided into visible (registered) and hidden parts *R* = *R*^(*v*)^ + *R*^(*h*)^. Infection and removal rates 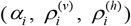 and the visibility coefficient *β*_*i*_ are supposed to be constant for every epidemic wave, i.e. for the time periods: 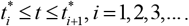 Summarizing eqs. (S1)-(S5) yields zero value of the derivative *d* (*S* + *I* ^(*v*)^ + *I* ^(*h*)^ + *R*^(*v*)^ + *R*^(*h*)^) / *dt*. Then the sum:

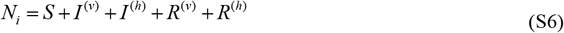

must be constant for every epidemic wave. We will consider the value *N*_*i*_ to be an unknown parameter of the model corresponding to the *i-th* wave, which is not equal to the known volume of population and must be estimated by observations. There is no need to assume that before the outbreak all people are susceptible, since many of them are protected by their immunity, distance, lockdowns, etc. Taking into account (S6), the initial conditions for the set of equations (S1)–(S5) at the beginning of every epidemic wave 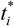 can be written as follows:

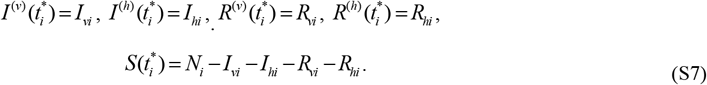

If at moment 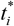 all previously infected persons are removed, we can take into account only cases starting to appear during *i-th* wave and use the initial conditions:

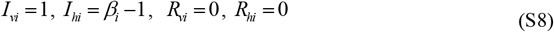

For the function ^*V(v)*^ = *I* ^*(v)*^ + *R*^*(v)*^ corresponding to the accumulated numbers of visible cases, eqs. (S2) and (S4) yield:

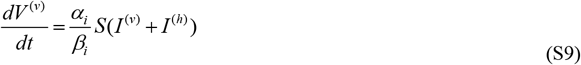

Eq. (S9) coincides with (3) and after dividing by (S1) and taking into account initial conditions (S7) allows us to obtain:

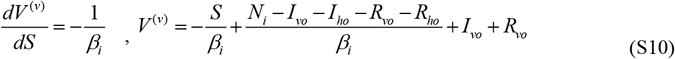

After putting in (S10) the initial conditions (S8), the first equation (2) can be obtained. For the general initial conditions (S7), eq. (2) is approximate.

### S2. Simulations of the first and second waves of pertussis epidemic in England in 2023 and 2024, [17]

To identify ten unknown parameters of the general model (S1)-(S7), differential equations have to be integrated numerically. The theoretical numbers of visible cases have to be compared with the results of observations in order to find the optimal values of parameters, which provide the best fitting.

To reduce the numbers of unknown parameters in the initial conditions (S7), sequential simulations of epidemic waves [19, 31] or (S8) can be used [17]. In the case of totally visible epidemic (*β* = 1) the exact analytical solution exists [19, 32-34], which in the case of initial conditions (S8) depends only on four unknown parameters *N*_*i*_, *α*_*i*_, 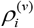and 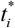. Since only two of these parameters are independent, the parameter identification procedure can be reduced to the problem of searching the maximum the correlation coefficient *r* (a function of these parameters and results of observations) [19, 32-34].

Two waves of the pertussis epidemic in England [18, 20] were simulated in [17] for the case *β*_1_ = *β*_2_ = 1, initial conditions (S8) and the results of observations listed in Table 1 (j=3-9 for the first wave and j=11-18 for the second one). The calculated optimal values of parameters are:

N_i_=50739.992 (3657890.47292358);

*α*_*i*_ =1.50488802805402e-05 (2.11266650706657e-06) [day]^-1^;

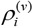 =0.752503596035381 (7.7088126606047) [day]^-1^;

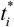=89.1566573802040 (333.429529143536) days;

(figures in brackets correspond to the second wave). These values were used to calculate the numbers of infectious *I(t)* and reproduction rates (1) shown in Fig. 1 by solid and dashed lines.

Since for the totally visible epidemic the average time of spreading the infection can be estimated as 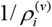 [8, 19] we can use the formula, [8]:

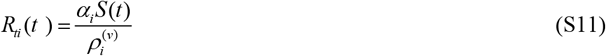

to calculate the effective reproduction numbers. Dotted curves in Fig.1 represent the results for the first (blue) and second (red) pertussis waves.

